# Genetic liability for endometriosis associates with pain, inflammatory, and metabolic traits: a polygenic score analysis

**DOI:** 10.64898/2026.06.18.26355987

**Authors:** Juliane L Beauchamp, Karina Banasik, Lisette J Kogelman, David Westergaard, Dea F Skipper, Olafur Davidsson, Sisse R Ostrowski, Christina Mikkelsen, Christian Erikstrup, Mie T Bruun, Bitten Aagaard, Ole B Pedersen, Erik Sørensen, Henrik Ullum, DBDS Genomic Cosnoritum, Palle D Rohde, Mette Nyegaard, Thomas F Hansen, Henriette S Nielsen

## Abstract

**Purpose:** Endometriosis is associated with pain, cardiometabolic, psychiatric, and immune comorbidities. We tested whether the genetic liability captured by an endometriosis polygenic risk score (PRS) extends to these comorbidities through shared pathways or operates independently of the disease.

**Methods:** In 168,238 women and 155,304 men of European ancestry from two Danish genetic studies linked to national health registers, we constructed an endometriosis PRS using LDpred2 from an independent GWAS meta-analysis (23,112 cases, 429,677 controls). Entropy balancing with doubly robust adjustment addressed selection bias. Comorbidity analyses were performed in both sexes to distinguish shared from endometriosis-specific pathways.

**Results:** The PRS was associated with endometriosis (OR 1.55 per SD, 95% CI 1.50–1.61; AUC 0.73) and with 20 of 29 comorbidities in women. Pain conditions (fibromyalgia, migraine, chronic back pain) and cardiometabolic conditions replicated in men, indicating shared pathways, whereas immune-mediated conditions associated in women only. Seventeen associations persisted in women without diagnosed endometriosis. High genetic risk was associated with increased healthcare utilisation and all-cause mortality (HR 1.05, 95% CI 1.01–1.10).

**Conclusion:** The endometriosis PRS captures a pain-inflammatory-metabolic genetic axis operating in both sexes, indicating that the genetic liability extends beyond the uterine disease and providing a rationale for targeted investigation and risk stratification.

## Introduction

Endometriosis is a chronic inflammatory disease defined by the growth of ectopic endometriotic lesions outside the uterine cavity^1^, affecting approximately 10% of women of reproductive age although prevalence varies depending on diagnostic thresholds and population characteristics^2^. Increasingly recognised as a systemic condition^3^, endometriosis is associated with a broad range of comorbidities beyond the reproductive system, including chronic pain conditions, cardiometabolic disease, and psychiatric disorders^4–8^. Women diagnosed with endometriosis demonstrate increased healthcare utilisation even prior to diagnosis^5,9^, and the disease is linked to reduced quality of life^10,11^ and considerable socioeconomic consequences^11–13^. Endometriosis has been identified as a major contributor to the women’s health gap, with effective management estimated to reclaim 250,000 disability-adjusted life years and generate an additional 12 billion USD in global GDP annually^14^. Prospective studies have linked endometriosis to increased risk of coronary heart disease^15^ and stroke, suggesting that the disease burden extends to cardiometabolic outcomes.

Endometriosis has a substantial genetic component^16^, and genome-wide association studies have identified shared risk loci between endometriosis and pain, inflammatory, and metabolic conditions^17^. Polygenic risk scores (PRS) quantify an individual’s genetic predisposition based on common risk variants and have demonstrated utility for risk prediction in other complex diseases^18^. Whether the genetic risk captured by an endometriosis PRS extends to pain-processing, inflammatory, and cardiometabolic pathways, and whether such associations operate independently of endometriosis itself, remains unknown.

In this study, we characterise the association of an endometriosis PRS with the disease itself, 29 comorbidities, and healthcare utilisation in 168,238 Danish women and 155,304 men and use sex-stratified analyses to distinguish endometriosis-specific from shared systemic genetic effects.

## Materials and Methods

### Study design

This registry-based cohort study analysed diagnoses and healthcare utilisation among genotyped participants of two Danish genetic studies linked to national health registers until 7 November 2024. The study period covers hospitalisations from 1977 onwards, prescriptions from 1994 onwards, and births from 1973 onwards.

### Study cohort

The study cohort comprised participants from the Copenhagen Hospital Biobank on Oral-Cardio-Metabolic Health (CHB-OCMS) and the Danish Blood Donor Study (DBDS). The CHB-OCMS is based on residual EDTA whole blood samples from patients aged >18 years treated at hospitals in the Capital Region of Denmark since 2009^19^. The DBDS consists of blood samples from volunteer blood donors recruited nationwide^20^. After genetic quality control and restriction to European ancestry, the analytical cohort comprised 323,542 individuals (236,145 from CHB-OCMS, 87,397 from DBDS; 168,238 women, 155,304 men).

### Data sources

Medical information was obtained from the Danish National Patient Registry (hospitalisations from public hospitals since 1977, private hospitals since 2002)^21^, the Danish National Prescription Registry (all dispensed prescriptions since 1994)^22^, and the Medical Birth Registry (all deliveries since 1973)^23^. Data linkage used the unique personal identification number assigned to all Danish residents via the Central Person Registry.

### Genotyping

Genotyping was conducted at deCODE Genetics using the Illumina Global Screening Array, as described elsewhere^20^. Imputation was performed using deCODE’s in-house pipeline^24^ based on a haplotype reference panel of WGS individuals from several cohorts^25^, including 8,671 Danish individuals^26^. We excluded samples whose principal components deviated >5 SD from the centroid of individuals with Danish-born parents, based on principal component analysis using FlashPCA2.0^27^ applied to quality-controlled, linkage disequilibrium (LD)-pruned genotype variants. Approximately 15% of the initial cohort was excluded due to non-European genetic ancestry.

### Exposure: polygenic risk score

The endometriosis PRS was calculated using LDpred2^28^ with the auto model. Summary statistics were derived from a large-scale GWAS meta-analysis comprising 23,112 cases and 429,677 controls^29^. These figures exclude the 23andMe cohort (37,182 cases, 251,255 controls) owing to data access restrictions, and the DBDS cohort (380 cases, 20,994 controls) to avoid sample overlap. No Danish samples were included in the discovery GWAS, ensuring no overlap between the GWAS and the target cohort. The LD matrix was based on 20,000 unrelated, randomly selected samples of European ancestry from the CHB-OCMS cohort, as LD structure is determined by ancestry rather than ascertainment.

The PRS was standardised to mean zero and unit variance within sex strata among European-ancestry participants. For comorbidity analyses, the PRS was rescaled so that one unit corresponds to a doubling of endometriosis risk, by dividing by ln(OR_endo_)/ln(2), where OR_endo_ is the per-SD odds ratio for endometriosis (1.55). This rescaling facilitates comparison with the endometriosis diagnosis effect; per-SD odds ratios are provided in Supplementary Table S1.

### Outcomes

#### Endometriosis and adenomyosis

Endometriosis and adenomyosis (endometrium infiltrating the myometrium) were identified from primary and secondary hospital diagnoses using ICD-8 code 625.3 and ICD-10 codes N80.0–N80.9. Adenomyosis is coded as N80.0 (“endometriosis of uterus”) in the Danish ICD-10 implementation and is therefore included in the overall endometriosis definition; adenomyosis was additionally analysed as a separate outcome using N80.0 alone.

#### Comorbidities

Comorbidities were identified through literature searches, resulting in 35 pre-specified conditions, of which 29 had sufficient case counts for analysis. Conditions were retained based on: (1) consistent associations reported across studies, (2) a minimum prevalence of 500 cases among women in the study cohort, and (3) documentation in the Danish health registries during the study period. Diagnoses were defined using the International Classification of Diseases, 8th revision (1977–1994) and 10th revision (1994 onwards), with ICD-8 codes mapped to ICD-10 equivalents using standardised conversion protocols^30^. Complete outcome definitions are provided in Supplementary Table S2.

#### Healthcare utilisation

Healthcare utilisation was measured as total hospital admissions, distinct ICD diagnoses, total dispensed prescriptions, pain prescriptions (Anatomical Therapeutic Chemical (ATC) group N02), and distinct ATC anatomical groups, each recorded across the full observation period. Prescription medications were classified using the ATC classification system. Observation time was calculated separately for each register: from the later of birth or registry start date to the earliest of emigration, death, or 7 November 2024.

### Statistical analysis

Cohort characteristics were summarised as medians with interquartile ranges for continuous variables and counts with percentages for categorical variables, stratified by endometriosis diagnosis and sex (Table 1).

**Table 1.** Baseline characteristics of the IPTW-weighted study cohort stratified by endometriosis diagnosis and sex, presented as median (IQR) for continuous variables and n (%) for categorical variables. CHB = Copenhagen Hospital Biobank; DBDS = Danish Blood Donor Study; DNPR = Danish National Patient Registry; PRS = polygenic risk score.

| Age (years) | 64 (46–79) | 50 (41–62) | 63 (45–78) | 66 (52–77) |
| --- | --- | --- | --- | --- |
| Year of birth | 1,959 (1,943–1,979) | 1,974 (1,961–1,983) | 1,960 (1,943–1,979) | 1,956 (1,944–1,972) |
| **Sub cohort** |  |  |  |  |
| CHB-OCMS | 122,161 (74%) | 3,280 (82%) | 125,441 (75%) | 110,704 (71%) |
| DBDS | 42,088 (26%) | 709 (18%) | 42,797 (25%) | 44,600 (29%) |
| Follow-up, DNPR (years) | 46.1 (38.9–47.8) | 47.1 (39.9–47.8) | 46.1 (39.0–47.8) | 47.8 (39.9–47.8) |
| Follow-up, prescriptions (years) | 30.9 (28.3–30.9) | 30.9 (30.9–30.9) | 30.9 (28.4–30.9) | 30.9 (26.6–30.9) |
| Parity |  |  |  |  |
| 1 | 28,449 (32%) | 947 (40%) | 29,396 (33%) |  |
| 2 | 37,928 (43%) | 981 (42%) | 38,909 (43%) |  |
| 3+ | 21,306 (24%) | 417 (18%) | 21,723 (24%) |  |
| Age at first child (years) | 27.7 (24.5–31.0) | 28.4 (24.4–32.2) | 27.7 (24.5–31.0) |  |
| Age at endometriosis (years) | — | 34 (28–41) | 34 (28–41) |  |
| Hospital admissions | 9 (4–17) | 17 (10–31) | 9 (5–17) | 3 (1–8) |
| Hospital admissions, grouped |  |  |  |  |
| 0-3 | 16,132 (10%) | 75 (1.9%) | 16,207 (10.0%) | 39,947 (34%) |
| 4-10 | 63,808 (40%) | 913 (23%) | 64,721 (40%) | 47,570 (40%) |
| 11-20 | 44,147 (28%) | 1,284 (32%) | 45,431 (28%) | 18,642 (16%) |
| 21-50 | 27,885 (18%) | 1,280 (32%) | 29,165 (18%) | 9,923 (8.4%) |
| 50+ | 6,541 (4.1%) | 437 (11%) | 6,978 (4.3%) | 2,227 (1.9%) |
| Endometriosis-related admissions | 0 (0–0) | 26 (14–48) | 0 (0–0) |  |
| Distinct ICD diagnoses | 6 (3–9) | 10 (6–14) | 6 (3–9) | 2 (1–3) |
| Distinct ICD diagnoses, grouped |  |  |  |  |
| 0-2 | 11,484 (7.2%) | 43 (1.1%) | 11,527 (7.1%) | 37,038 (31%) |
| 3-5 | 46,669 (29%) | 541 (14%) | 47,210 (29%) | 60,991 (52%) |
| 6-10 | 62,620 (40%) | 1,358 (34%) | 63,978 (39%) | 18,921 (16%) |
| 11-20 | 35,729 (23%) | 1,809 (45%) | 37,538 (23%) | 1,356 (1.1%) |
| 20+ | 2,011 (1.3%) | 238 (6.0%) | 2,249 (1.4%) | 3 (<0.1%) |
| Prescriptions | 181 (91–433) | 176 (108–311) | 181 (92–430) | 137 (44–351) |
| Prescriptions per year | 7 (4–17) | 7 (5–12) | 7 (4–17) | 7 (3–15) |
| Pain prescriptions (N02) | 7 (1–35) | 7 (1–25) | 7 (1–35) | 5 (1–20) |
| Pain prescriptions per year | 1.8 (1.0–4.0) | 1.5 (1.0–3.0) | 1.8 (1.0–4.0) | 1.5 (1.0–3.3) |
| Distinct ATC codes | 36 (24–52) | 40 (29–55) | 36 (24–52) | 27 (16–42) |
| Distinct ATC anatomical groups | 10 (8–11) | 10 (9–11) | 10 (8–11) | 9 (7–10) |
| PRS (standardised) | -0.01 (-0.68–0.66) | 0.41 (-0.26–1.11) | 0.00 (-0.67–0.68) | 0.00 (-0.68–0.68) |

As previous studies have found the sub-cohorts CHB-OCMS and DBDS to differ in baseline morbidity and demographic composition^31^, we applied inverse probability of treatment weighting (IPTW) via entropy balancing^32^ to mitigate selection bias induced by the differing recruitment mechanisms of CHB-OCMS and DBDS. The weighting model balanced means, variances, and pairwise interactions of year of birth, sex, a birth-before-1977 indicator, and five genetic principal components (PCs) between CHB-OCMS and DBDS, targeting the DBDS covariate distribution as the reference population; weights were trimmed at the 97.5th percentile. Covariate balance is provided in Supplementary Table S3. All subsequent analyses were conducted on the IPTW-weighted sample with robust standard errors and additional covariate adjustment, providing doubly robust inference^33,34^. All regression models were adjusted for year of birth modelled with natural cubic splines, an indicator for birth before 1977 (i.e. left censoring), and the first five genetic PCs.

Adjusted logistic regression was used to estimate per-SD odds ratios for endometriosis and adenomyosis among women. Additionally, odds ratios were estimated per PRS decile, with the 5th decile as reference. The discriminative ability of the PRS was evaluated using the area under the receiver operating characteristic curve (AUC) for a covariate-only and a PRS-augmented model, with improvement tested by likelihood ratio and DeLong tests. Optimism-corrected C-statistics were obtained via 200 paired bootstrap resamples. Time-dependent cumulative/dynamic AUC was computed at ages 20, 30, 40, 50, and 60 years to assess how discrimination varies across the age span.

For comorbidity analyses, the PRS was rescaled so that one unit corresponds to a doubling of endometriosis risk (see Exposure). Logistic regression was used to estimate odds ratios for 29 comorbidities. Two exposures were examined: the PRS on the doubling scale and endometriosis diagnosis, the latter modelled as both any-time co-occurrence and temporally ordered (endometriosis preceding the comorbidity). Analyses were performed among all women and among women without endometriosis. To investigate whether the PRS captures shared biological pathways not dependent on uterine endometriosis, the same comorbidity analyses were performed in men; associations present in men would indicate pleiotropy rather than mediation through endometriotic disease. We adjusted for multiple testing using Benjamini–Hochberg false discovery rate correction applied to the 29 comorbidity tests within each analysis stratum. Age-as-time-scale Cox proportional hazards models were fitted as supplementary analyses; logistic regression was preferred as the primary approach because proportional hazards did not hold for the majority of comorbidities, making the lifetime odds ratio a more stable summary measure.

To assess healthcare burden, individuals were stratified into low-risk (PRS deciles 1–3), average-risk (deciles 4–7), and high-risk (deciles 8–10) groups. Quasi-Poisson regression with an offset for log observation time was used to estimate incidence rate ratios for healthcare utilisation between risk groups. All-cause mortality was compared across the same risk groups using Cox proportional hazards regression.

As sensitivity analyses, all primary analyses were repeated without IPTW (covariate adjustment only) to assess the impact of selection bias correction, and a second set of weights was estimated from a selection model that additionally included the PRS to assess exposure-dependent selection. Decile and utilisation analyses were not corrected for multiple testing and should be interpreted as exploratory.

All analyses were conducted using R v4.5.0. Analytical code is available at [gitlab URL]. This study is reported in accordance with the STROBE guidelines for observational studies^35^.

### Ethics

The CHB-OCMS and DBDS have been approved by the Zealand Regional and National Committees on Health Research Ethics (SJ-989 and NVK-1900988) and the Danish Data Protection Agency (P-2022-913 and P-2019-99)

### Data availability

Individual-level data cannot be shared owing to Danish data protection legislation. Summary-level results and analytical code are available at [gitlab URL].

## Results

### Cohort characteristics

The analytical cohort comprised 168,238 women (median age 63 years, IQR 45–78) and 155,304 men (median age 66 years, IQR 51–77) of European ancestry, of whom 236,145 were from CHB-OCMS and 87,397 from DBDS. Among women, 3,989 (2.37%) had a hospital diagnosis of endometriosis and 1,584 (0.94%) had adenomyosis. Entropy balancing achieved covariate balance between CHB-OCMS and DBDS (all standardised mean differences <0.05, all variance ratios <2.0; Supplementary Table S3). Cohort characteristics are presented in Table 1.

### PRS and endometriosis

The PRS was associated with endometriosis (OR 1.55 per SD increase, 95% CI 1.50–1.61, p<0.001) and adenomyosis (OR 1.48, 95% CI 1.38–1.59, p<0.001) (Table 2). When stratified by PRS decile, women in the tenth decile had an endometriosis prevalence of 4.7% compared with 1.1% in the first decile (Figure 1A, 1C).

**Figure 1.**
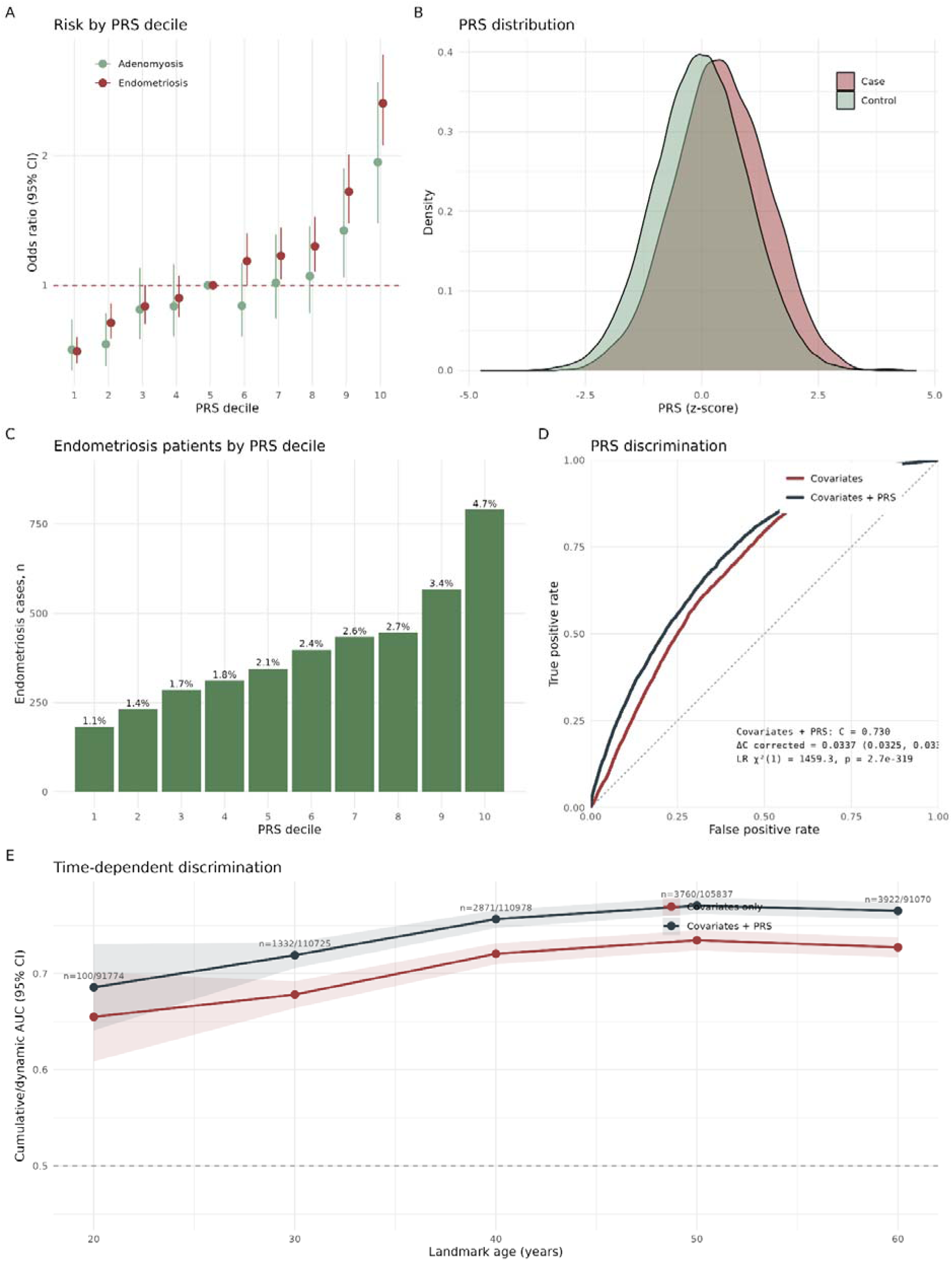
Polygenic risk score (PRS) for endometriosis: distribution, predictive performance, and risk stratification. **(A)** Odds ratios with 95% confidence intervals for endometriosis and adenomyosis across PRS deciles, with the 5th decile as reference. **(B)** PRS distribution among endometriosis cases and controls. **(C)** Number of endometriosis cases per PRS decile; percentages indicate prevalence within each decile. **(D)** Receiver operating characteristic curves for covariate-only and PRS-augmented models (AUC 0.697 vs 0.730; delta-C 0.034). **(E)** Time-dependent cumulative/dynamic AUC at landmark ages 20, 30, 40, 50, and 60 years with 95% DeLong confidence intervals; annotations show cases/controls at each age. All analyses conducted on the IPTW-weighted sample.

**Table 2.** Per-standard deviation odds ratios (OR) for endometriosis and adenomyosis. Adjusted for year of birth (natural cubic splines), birth before 1977, and five genetic principal components. IPTW-weighted with HC1 sandwich standard errors.

| Outcome | N | Cases | OR (95% CI) | P-value |
| --- | --- | --- | --- | --- |
| Endometriosis | 168,238 | 3,989 | 1.55 (1.50–1.61) | 2.5e-125 |
| Adenomyosis | 168,238 | 1,584 | 1.48 (1.38–1.59) | 7.4e-28 |

The PRS-augmented model achieved an AUC of 0.730 (95% CI 0.723–0.737) compared with 0.697 (95% CI 0.690–0.704) for covariates alone, an improvement of 0.034 (95% CI 0.033–0.034) supported by both likelihood ratio (chi-squared 1459, df=1, p<0.001) and DeLong tests (p<0.001) (Figure 1D). Optimism-corrected C-statistics (200 paired bootstrap resamples) were 0.729 and 0.695, respectively. Time-dependent AUC increased from 0.69 (95% CI 0.64–0.73) at age 20 to 0.77 (95% CI 0.76–0.78) at age 50, plateauing thereafter (Figure 1E).

### Comorbidity associations

The PRS was associated with 20 of 29 analysed comorbidities in women after FDR correction (Table 4, Figure 2). Comorbidity ORs are reported on the doubling scale (see Methods).

**Figure 2.**
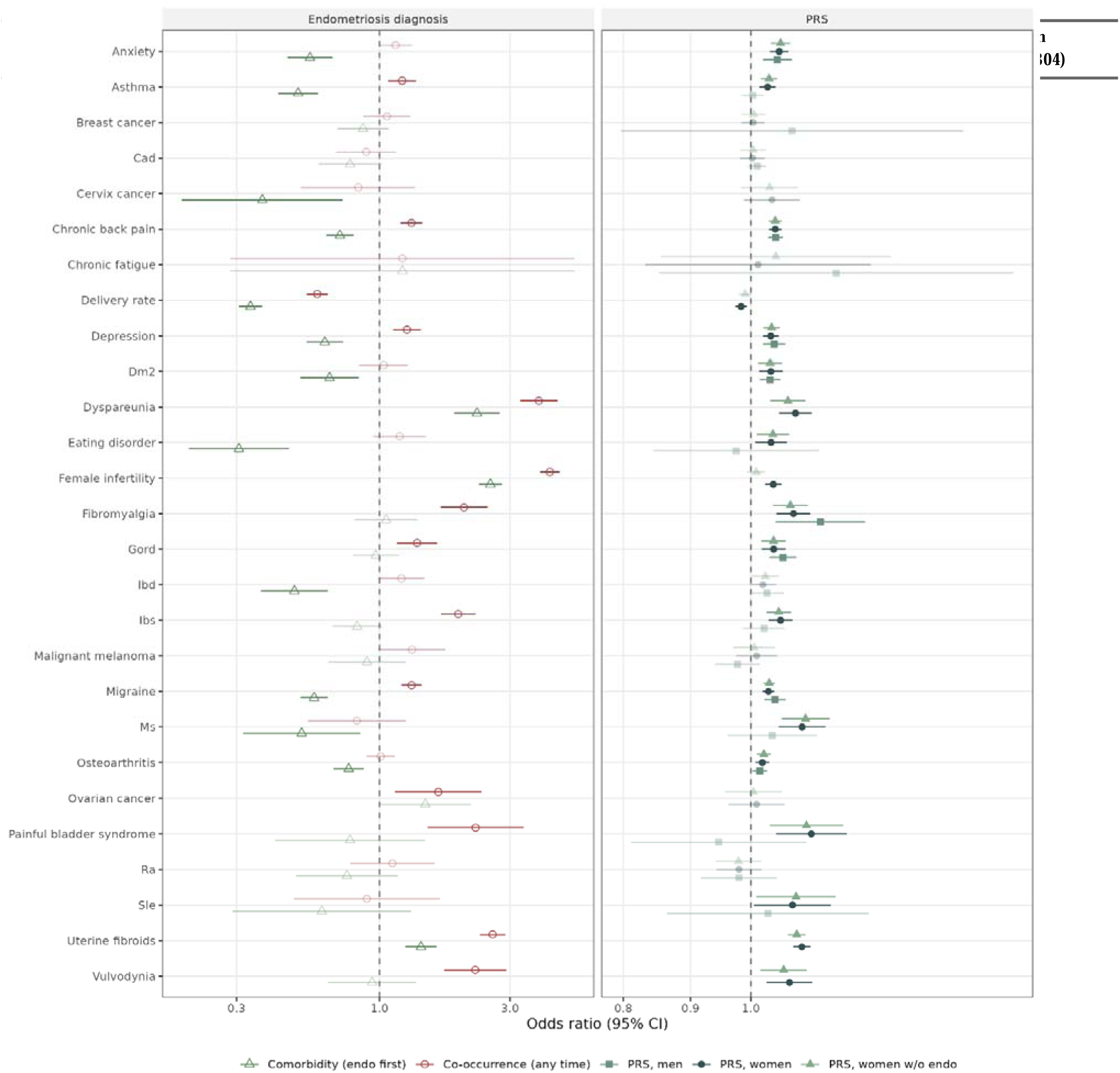
Forest plot of odds ratios for the association between the endometriosis polygenic risk score (PRS; doubling scale) and 29 comorbidities, and between endometriosis diagnosis and the same comorbidities. Left panel: endometriosis diagnosis as exposure (co-occurrence regardless of temporal order; comorbidity with endometriosis preceding). Right panel: PRS as exposure (all women; women without (w/o) endometriosis; men). Faded points indicate BH-FDR >= 0.05. All analyses conducted on the IPTW-weighted sample with robust standard errors.

All eight tested pain-related conditions were associated with the PRS in women (Table 4, Figure 2), with the strongest associations for fibromyalgia (OR 1.08, 95% CI 1.05–1.11) and dyspareunia (OR 1.08, 95% CI 1.05–1.11). Fibromyalgia (OR 1.13, 95% CI 1.04–1.22), chronic back pain (OR 1.04, 95% CI 1.03–1.06), migraine (OR 1.04, 95% CI 1.02–1.06), and osteoarthritis (OR 1.02, 95% CI 1.00–1.03) replicated in men. Among gynaecological outcomes (women only), uterine fibroids showed the strongest association (OR 1.09, 95% CI 1.08–1.11).

The PRS was associated with type 2 diabetes mellitus and gastro-oesophageal reflux in both sexes, as well as anxiety (women OR 1.05, 95% CI 1.03–1.07; men OR 1.05, 95% CI 1.02–1.07) and depression (women OR 1.04, 95% CI 1.02–1.05; men OR 1.04, 95% CI 1.02–1.06); in women only, associations were observed with multiple sclerosis (OR 1.09, 95% CI 1.05–1.14), asthma (OR 1.03, 95% CI 1.02–1.04), and SLE (OR 1.08, 95% CI 1.01–1.15) (Table 4).

Endometriosis diagnosis was associated with 15 of 29 comorbidities (any-time co-occurrence) and 17 of 29 (temporally ordered). Several conditions associated with the PRS showed no association with the diagnosis variable (Figure 2). Among women without an endometriosis diagnosis, 17 of 29 comorbidities remained associated with the PRS.

### Healthcare burden

High-risk carriers (PRS deciles 8–10) had more hospital admissions (mean 15.1 [high] vs 14.4 [average] vs 13.8 [low]), more distinct ICD diagnoses (6.9 vs 6.6 vs 6.4), and more dispensed prescriptions (356 vs 343 vs 327) than average- and low-risk groups, even when accounting for differences in follow-up time (Table 3). These differences persisted among women without an endometriosis diagnosis and were also observed in men. Furthermore, men and women with high PRS had the highest rates of analgesic and pain-related medication use, reflected in elevated prescription rates within ATC groups N (nervous system) and M (musculoskeletal system) (Supplementary Table S4). In the pooled cohort, women and men, all-cause mortality was higher in the high-risk compared with the low-risk group (HR 1.05, 95% CI 1.01–1.10, p=0.008); no difference was observed between adjacent risk groups.

**Table 3.**
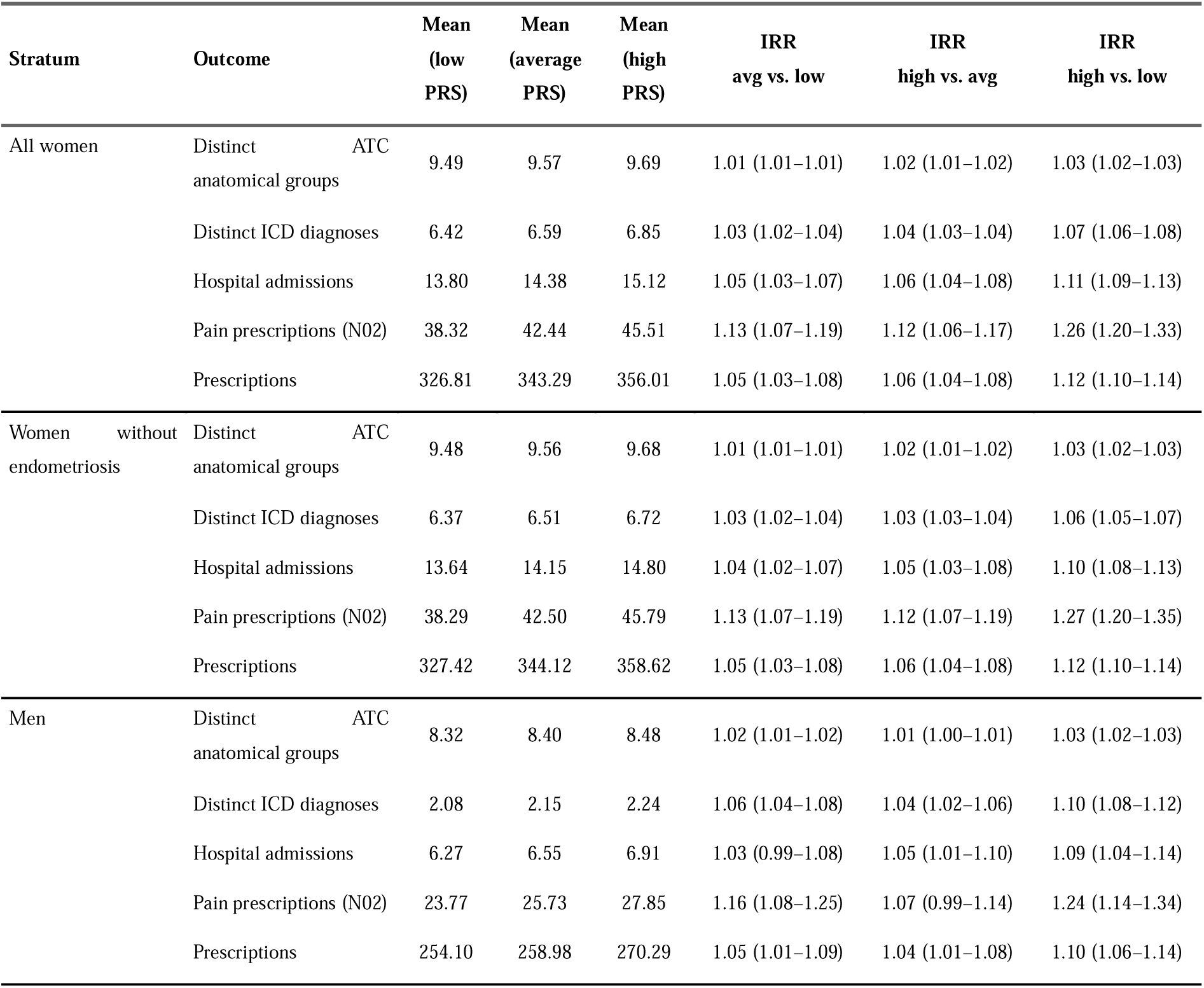
Healthcare utilisation by polygenic risk score (PRS) risk group (low, deciles 1–3; average, deciles 4–7; high, deciles 8–10). Incidence rate ratios from quasi-Poisson regression with offset for log observation time, IPTW-weighted. Presented for all women, women without endometriosis, and men.

**Table 4.** Association between the endometriosis PRS (doubling scale) and 29 comorbidities. Odds ratios with 95% confidence intervals from logistic regression, IPTW-weighted with HC1 sandwich standard errors. Results for: PRS in all women, PRS in women without endometriosis, PRS in men, and endometriosis diagnosis (any-time co-occurrence and temporally ordered). Asterisks indicate BH-FDR significance (* <0.05, ** <0.01, *** <0.001).

| Phenotype | OR, PRS<br>(women) | OR, PRS<br>(women w/o endo) | OR, PRS<br>(men) | OR, endo<br>(any time) | OR, endo<br>(before) |
| --- | --- | --- | --- | --- | --- |
| Anxiety | 1.05 (1.03–1.07)*** | 1.05 (1.04–1.07)*** | 1.05 (1.02–1.07)** | 1.14 (0.99–1.32) | 0.56 (0.46–0.67)*** |
| Asthma | 1.03 (1.02–1.04)*** | 1.03 (1.02–1.05)*** | 1.00 (0.98–1.02) | 1.21 (1.07–1.36)** | 0.50 (0.43–0.60)*** |
| Breast cancer | 1.00 (0.98–1.02) | 1.00 (0.98–1.03) | 1.07 (0.80–1.45) | 1.06 (0.87–1.30) | 0.87 (0.70–1.08) |
| Coronary artery disease | 1.00 (0.98–1.02) | 1.00 (0.98–1.03) | 1.01 (1.00–1.03) | 0.89 (0.70–1.15) | 0.78 (0.60–1.02) |
| Cervix cancer | 1.04 (0.99–1.09) | 1.03 (0.98–1.09) | — | 0.84 (0.52–1.35) | 0.37 (0.19–0.73)** |
| Chronic back pain | 1.04 (1.03–1.06)*** | 1.04 (1.03–1.06)*** | 1.04 (1.03–1.06)*** | 1.31 (1.19–1.44)*** | 0.72 (0.64–0.80)*** |
| Chronic fatigue | 1.01 (0.83–1.23) | 1.04 (0.85–1.28) | 1.16 (0.85–1.58) | 1.21 (0.28–5.15) | 1.21 (0.29–5.16) |
| Delivery rate | 0.98 (0.97–0.99)** | 0.99 (0.98–1.00) | — | 0.59 (0.54–0.65)*** | 0.34 (0.31–0.37)*** |
| Depression | 1.04 (1.02–1.05)*** | 1.04 (1.02–1.05)*** | 1.04 (1.02–1.06)*** | 1.26 (1.12–1.41)*** | 0.63 (0.54–0.74)*** |
| Type 2 diabetes mellitus | 1.04 (1.01–1.06)** | 1.03 (1.01–1.06)** | 1.03 (1.02–1.05)*** | 1.03 (0.84–1.27) | 0.66 (0.51–0.84)** |
| Dyspareunia | 1.08 (1.05–1.11)*** | 1.07 (1.03–1.10)*** | — | 3.82 (3.27–4.47)*** | 2.27 (1.88–2.75)*** |
| Eating disorder | 1.04 (1.01–1.06)* | 1.04 (1.01–1.07)* | 0.97 (0.84–1.13) | 1.18 (0.95–1.48) | 0.31 (0.20–0.47)*** |
| Female infertility | 1.04 (1.03–1.06)*** | 1.01 (0.99–1.02) | — | 4.19 (3.86–4.56)*** | 2.54 (2.31–2.80)*** |
| Fibromyalgia | 1.08 (1.05–1.11)*** | 1.07 (1.04–1.10)*** | 1.13 (1.04–1.22)** | 2.04 (1.67–2.48)*** | 1.06 (0.81–1.37) |
| Gastritis duodenitis | 0.73 (0.64–0.83)*** | — | — | — | — |
| Gastro-oesophageal<br>reflux disease | 1.04 (1.02–1.06)*** | 1.04 (1.02–1.06)*** | 1.06 (1.03–1.08)*** | 1.37 (1.16–1.62)*** | 0.97 (0.80–1.18) |
| Haemorrhoids | 1.11 (1.01–1.23) | 1.09 (0.98–1.20) | — | 1.37 (0.57–3.26) | 0.00 (0.00–0.00)*** |
| Inflammatory bowel disease | 1.02 (1.00–1.05) | 1.03 (1.00–1.05) | 1.03 (1.00–1.06) | 1.20 (0.99–1.46) | 0.49 (0.37–0.65)*** |
| Irritable bowel syndrome | 1.05 (1.03–1.08)*** | 1.05 (1.03–1.07)*** | 1.02 (0.99–1.06) | 1.94 (1.68–2.24)*** | 0.83 (0.67–1.02) |
| Malignant melanoma | 1.01 (0.97–1.05) | 1.01 (0.97–1.04) | 0.98 (0.94–1.02) | 1.31 (0.99–1.75) | 0.90 (0.65–1.25) |
| Migraine | 1.03 (1.02–1.04)*** | 1.03 (1.02–1.04)*** | 1.04 (1.02–1.06)*** | 1.31 (1.20–1.42)*** | 0.58 (0.51–0.65)*** |
| Multiple sclerosis | 1.09 (1.05–1.14)*** | 1.10 (1.06–1.15)*** | 1.04 (0.96–1.12) | 0.83 (0.55–1.25) | 0.52 (0.32–0.85)* |
| Osteoarthritis | 1.02 (1.01–1.03)** | 1.02 (1.01–1.04)*** | 1.02 (1.00–1.03)* | 1.01 (0.90–1.14) | 0.77 (0.68–0.88)*** |
| Ovarian cancer | 1.01 (0.96–1.06) | 1.00 (0.96–1.06) | — | 1.64 (1.14–2.36)* | 1.47 (1.00–2.16) |
| Painful bladder syndrome | 1.11 (1.05–1.18)** | 1.10 (1.03–1.18)** | 0.95 (0.81–1.10) | 2.25 (1.50–3.36)*** | 0.78 (0.42–1.46) |
| Rheumatoid arthritis | 0.98 (0.94–1.02) | 0.98 (0.94–1.02) | 0.98 (0.92–1.05) | 1.11 (0.78–1.59) | 0.76 (0.49–1.17) |
| Systemic lupus erythematosus | 1.08 (1.01–1.15)* | 1.08 (1.01–1.16)* | 1.03 (0.86–1.23) | 0.90 (0.49–1.66) | 0.62 (0.29–1.30) |
| Uterine fibroids | 1.09 (1.08–1.11)*** | 1.08 (1.07–1.10)*** | — | 2.59 (2.33–2.88)*** | 1.42 (1.24–1.62)*** |
| Vulvodynia | 1.07 (1.03–1.11)** | 1.06 (1.02–1.10)* | 1.26 (0.94–1.71) | 2.24 (1.72–2.91)*** | 0.94 (0.65–1.36) |

## Discussion

In women, the endometriosis PRS was associated with the disease and with the majority of tested comorbidities, spanning pain, cardiometabolic, psychiatric, and immune-mediated conditions. The pattern of comorbidity associations was not uniform: pain and cardiometabolic conditions replicated in men, while immune-mediated conditions did not. This sex-stratified dissociation indicates that the PRS captures at least two distinct biological signals, a sex-independent pain-inflammatory-metabolic axis and sex-specific or endometriosis-mediated immune pathways, and that endometriosis may be one clinical manifestation of a broader genetic predisposition.

### PRS and endometriosis

Our findings align with previous studies reporting comparable effect sizes and modest discrimination for endometriosis PRS^36–38^. The higher AUC in this study reflects the inclusion of covariates in the prediction model; the incremental discrimination attributable to the PRS above covariates alone was modest but consistent. While Kloeve-Mogensen *et al*.^36^ did not identify an association with adenomyosis, our study detected one, likely due to increased statistical power. The discovery GWAS is enriched for surgically confirmed and severe cases, which may bias the PRS toward pathways active in advanced disease and partially explain the strong associations with pain conditions.

### A shared pain-inflammatory-metabolic genetic axis

The association of the endometriosis PRS with fibromyalgia, migraine, chronic back pain, and type 2 diabetes mellitus in men is the central mechanistic finding of this study. As endometriosis does not occur in men, these associations indicate shared genetic pathways operating independently of diagnosed endometriosis. The pain conditions replicating in both sexes overlap substantially with the clinical construct of chronic overlapping pain conditions, a cluster of disorders thought to share central sensitisation mechanisms. Consistent with this, individuals with high PRS in both sexes had the highest prescription rates within ATC groups N and M, indicating greater use of analgesics and pain-related medications. Our genetic findings provide evidence that this clinical co-occurrence has a heritable basis.

The cardiometabolic associations are consistent with prospective epidemiological evidence linking endometriosis to increased risk of coronary heart disease^15^ and with the genetic correlations reported by Rahmioglu et al.^17^ and the Mendelian randomisation analyses of McGrath et al.^39^. These converging lines of evidence suggest that endometriosis and cardiometabolic conditions share inflammatory or metabolic pathways with a genetic basis, rather than the cardiometabolic risk being a downstream consequence of endometriosis alone.

In contrast, immune-mediated conditions associated with the PRS in women were not associated in men. These conditions may involve endometriosis-specific pathways, such as oestrogen-dependent immune dysregulation, or sex-differentially regulated genetic effects. The dissociation between conditions that replicate across sexes and those that do not provides a framework for prioritising mechanistic investigation: the shared pain-metabolic axis is amenable to study in both sexes and in non-endometriosis populations.

### Comorbidity associations in context

PRS-comorbidity associations persisted after excluding women with diagnosed endometriosis, consistent with the findings of McGrath et al.^40^. However, given the substantial underdiagnosis in registry-based cohorts, these sensitivities analyses do not exclude associations mediated by undiagnosed endometriosis.

Exceptions were female infertility and delivery, where diagnosis showed stronger associations than the PRS. The infertility association likely reflects ascertainment bias, as endometriosis is frequently diagnosed during fertility workup. That the PRS does not associate strongly with reproductive outcomes, while associating with pain and metabolic conditions, reinforces the interpretation that the PRS captures a systemic inflammatory-metabolic predisposition rather than the anatomical or mechanical consequences of endometriotic lesions.

### Healthcare utilisation

Although utilisation was higher in high-PRS groups regardless of clinical diagnosis, these analyses were not adjusted for overall comorbidity burden. The increased utilisation may partly reflect the cumulative effect of PRS-associated comorbidities rather than an independent consequence of genetic risk. The mortality finding should be interpreted cautiously given that the lower confidence bound only narrowly excludes the null. Future analyses adjusting for comorbidity indices could clarify whether excess utilisation exists beyond what associated conditions explain.

### Strengths and limitations

This study benefits from a large sample size and the validated Danish national health registers enabling comprehensive, longitudinal phenotyping. No Danish samples were included in the discovery GWAS, eliminating sample overlap. The use of inverse probability weighting via entropy balancing addresses selection bias from combining hospital-based and population-based cohorts, and the sex-stratified analyses provide a principled test for distinguishing pleiotropy from endometriosis-mediated effects.

The primary comorbidity analysis assessed lifetime prevalence; temporal relationships cannot be firmly established. The observed endometriosis prevalence indicates substantial underdiagnosis, meaning that sensitivity analyses excluding diagnosed cases still leave most true cases in the comparison group. PRS-comorbidity associations described as “independent of endometriosis” should be interpreted as independent of diagnosed endometriosis.

The CHB-OCMS is enriched for patients with cardiometabolic diseases; the DBDS recruits healthy blood donors. Although entropy balancing achieved covariate balance, residual selection bias cannot be excluded. Further, the absence of primary care data may contribute to under-ascertainment. The large sample size provides sufficient statistical power to detect small effect sizes that may not be clinically meaningful at the individual level. The PRS was derived from European-ancestry populations, limiting generalizability beyond European ancestry.

## Conclusion

The endometriosis PRS captures a pain-inflammatory-metabolic genetic axis that operates in both sexes, indicating that the genetic liability underlying endometriosis extends beyond the ectopic endometrium. These findings reframe endometriosis-associated comorbidity as partly reflecting shared genetic architecture rather than disease sequelae alone, and identify specific pathophysiological processes, pain processing, inflammation, and cardiometabolic regulation, where mechanistic investigation and targeted therapeutic development are warranted. Prospective evaluation of PRS-based risk stratification may benefit not only endometriosis detection but also the identification and management of associated morbidity across the full spectrum of genetic risk.

## Declaration of generative AI and AI-assisted technologies in the manuscript preparation process

During the preparation of this work, the authors used large language models for improvement of language and proofreading. The authors reviewed and edited the output as needed and takes full responsibility for the content of the published article.

## Author contributions

JLB: conceptualization, formal analysis, data curation, writing – original draft. KB: conceptualization, data curation, methodology, writing – draft, review & editing. LJAK: methodology, supervision, writing – review & editing. DW: data curation, methodology, analysis, software, writing – review & editing. DFS: data curation, writing – review & editing. OD: genotyping, data curation. SRO: resources, data curation. OBP: resources, data curation. CM: resources, data curation. CE: resources, data curation. MTB: resources, data curation. BA: resources, data curation. ES: resources, data curation. HU: resources, writing – review & editing. VS: genotyping, data curation. PDR: methodology, writing – review & editing. MN: methodology, writing – review & editing. TFH: conceptualization, supervision, funding acquisition, writing – review & editing. HSN: conceptualization, supervision, AP acquisition, writing – review & editing. All authors reviewed and approved the final manuscript.

## Acknowledgements

We thank the participants of the Copenhagen Hospital Biobank and the Danish Blood Donor Study. We acknowledge the DBDS genomic consortium for their contributions to genotyping and data infrastructure.

---

**Supplementary Table S1.**
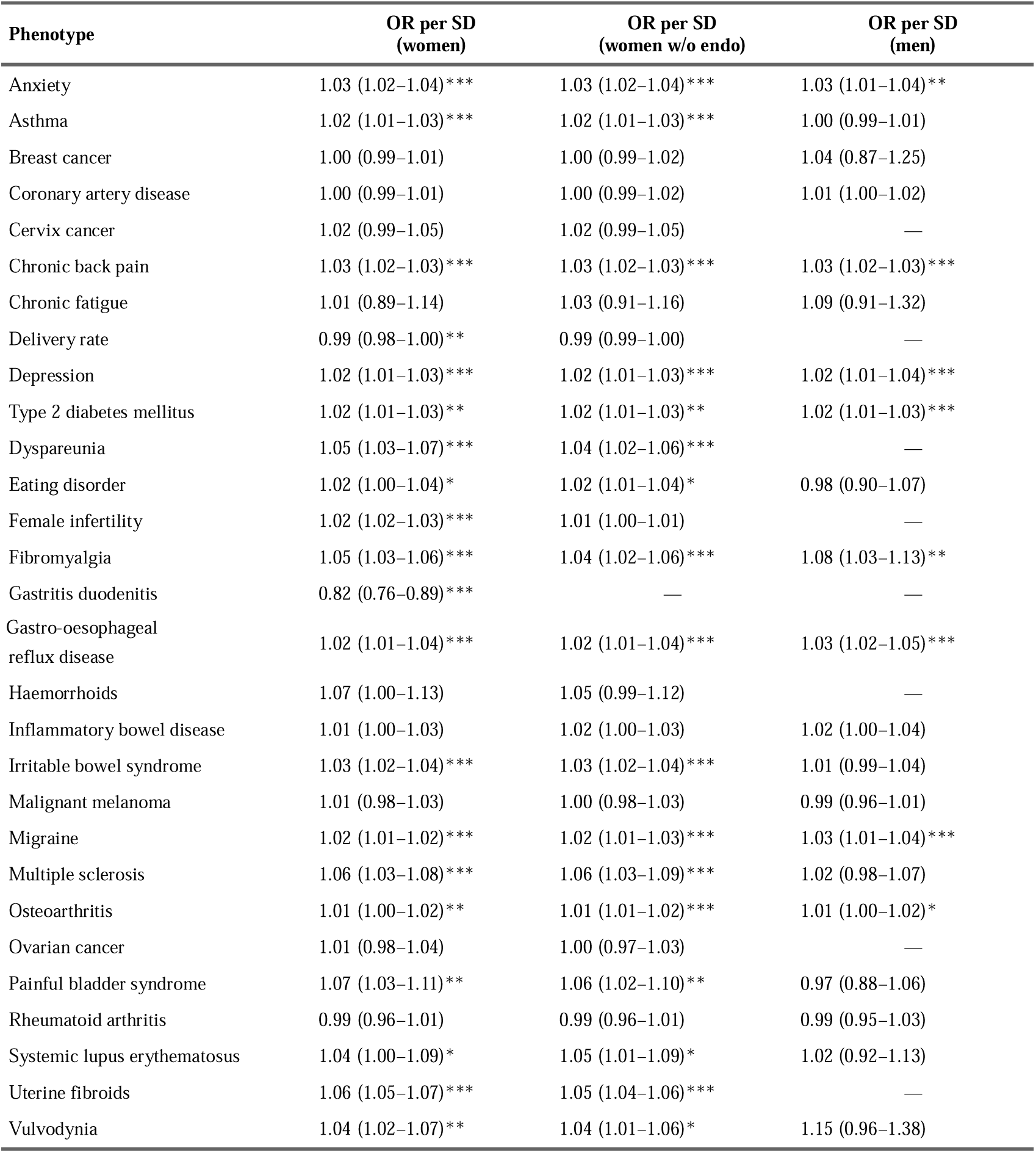
Per-SD odds ratios for associations between the endometriosis PRS and 29 comorbidities. Odds ratios (OR) per SD with 95% confidence intervals (CI) from logistic regression, IPTW-weighted with HC1 sandwich standard errors. Results for: PRS in all women, PRS in women without (w/o) endometriosis (endo), PRS in men, and endometriosis diagnosis (any-time co-occurrence and temporally ordered). Asterisks indicate BH-FDR significance (* <0.05, ** <0.01, *** <0.001). — Indicates that the logistic regression model did not converge due to insufficient case counts within this specific sub-stratum.

**Supplementary Table S2.** Outcome definitions. ICD-10 codes shown without the Danish ‘D’ prefix. * denotes wildcard matching all sub-codes.

| Category | # | Outcome | ICD-10 | ICD-8 | ATC |
| --- | --- | --- | --- | --- | --- |
| Reproductive outcomes | 1 | Uterine fibroids | D25* | 21899 | — |
|  | 2 | Delivery rate | O80*, O81*, O82*, O83*, O84* | 65*, 66* | — |
|  | 3 | Female infertility | N970*, N971*, N972*, N973*, N978*, N979* | 628* | — |
| Pain outcomes | 4 | Fibromyalgia | M797* | 71790 | — |
|  | 5 | Chronic fatigue | R53*, R688A9B1 | 79010, 79019 | — |
|  | 6 | Migraine | G43* | 3460 | N02C* |
|  | 7 | Chronic back pain | M54* | 7170, 72879, 72859, 72899 | — |
|  | 8 | Osteoarthritis | M15*, M16*, M17*, M18*, M19* | 7130* | — |
|  | 9 | Painful bladder syndrome | N301 | 59503 | — |
|  | 10 | Vulvodynia | N948A | 78679 | — |
|  | 11 | Dyspareunia | N941 | 78670 | — |
|  | 12 | Irritable bowel syndrome | K58* | 56419 | — |
| Immune-mediated outcomes | 13 | Asthma | J450, J451, J458, J459 | 4930* | — |
|  | 14 | Systemic lupus erythematosus | M32, M321, M328, M329 | 73419 | — |
|  | 15 | Rheumatoid arthritis | M05* | 71219, 71239 | — |
|  | 16 | Multiple sclerosis | G35* | 3400* | — |
|  | 17 | Inflammatory bowel disease | K50*, K51* | 5630*, 56319 | — |
| Gastro-intestinal and metabolic | 18 | Gastro-oesophageal reflux disease | K21* | — | — |
|  | 19 | Gastritis/duodenitis | K29* | 535* | — |
|  | 20 | Haemorrhoids | K640, K641, K642, K643, K644, K648, K649 | 455* | — |
|  | 21 | Type 2 diabetes mellitus | E11* | 2500* | — |
| Psychiatric outcomes | 22 | Anxiety | F40*, F41* | 30009, 30029 | — |
|  | 23 | Depression | F32*, F33* | 29609, 29809, 29629 | — |
|  | 24 | Eating disorder | F50* | 30650, 30658, 30659 | — |
| Malignant outcomes | 25 | Ovarian cancer | C56* | 1830* | — |
|  | 26 | Breast cancer | C50* | 1740* | — |
|  | 27 | Malignant melanoma | C43* | 172* | — |
|  | 28 | Cervix cancer | C53* | 1800* | — |
| Cardiovascular outcomes | 29 | Coronary artery disease | I21*, I22*, I23*, I24*, I25* | 41199, 41099, 41409, 41499, 41299, 41209, 41009, 41109 | — |

**Supplementary Table S3.**

| Model | Variable | Type | Diff. (unadj.) | VR (unadj.) | Diff. (adj.) | VR (adj.) | Mean balanced | Variance balanced |
| --- | --- | --- | --- | --- | --- | --- | --- | --- |
| **Primary** | Year of birth | Contin. | -1.485 | 2.08 | -0.019 | 1.01 | Balanced, <0.05 | Balanced, <2 |
|  | Sex (male) | Binary | -0.042 | — | -0.012 | — | Balanced, <0.05 | — |
|  | Born before 1977 | Binary | 0.280 | — | 0.014 | — | Balanced, <0.05 | — |
|  | PC1 | Contin. | -0.117 | 1.12 | -0.015 | 1.00 | Balanced, <0.05 | Balanced, <2 |
|  | PC2 | Contin. | 0.006 | 1.05 | 0.000 | 1.00 | Balanced, <0.05 | Balanced, <2 |
|  | PC3 | Contin. | -0.110 | 1.20 | -0.011 | 1.00 | Balanced, <0.05 | Balanced, <2 |
|  | PC4 | Contin. | -0.056 | 1.00 | -0.009 | 1.00 | Balanced, <0.05 | Balanced, <2 |
|  | PC5 | Contin. | -0.085 | 1.10 | -0.008 | 1.00 | Balanced, <0.05 | Balanced, <2 |
| **With PRS** | Year of birth | Contin. | -1.485 | 2.08 | -0.020 | 1.01 | Balanced, <0.05 | Balanced, <2 |
|  | Sex (male) | Binary | -0.042 | — | -0.012 | — | Balanced, <0.05 | — |
|  | Born before 1977 | Binary | 0.280 | — | 0.014 | — | Balanced, <0.05 | — |
|  | PC1 | Contin. | -0.117 | 1.12 | -0.015 | 1.00 | Balanced, <0.05 | Balanced, <2 |
|  | PC2 | Contin. | 0.006 | 1.05 | 0.000 | 1.00 | Balanced, <0.05 | Balanced, <2 |
|  | PC3 | Contin. | -0.110 | 1.20 | -0.012 | 1.00 | Balanced, <0.05 | Balanced, <2 |
|  | PC4 | Contin. | -0.056 | 1.00 | -0.009 | 1.00 | Balanced, <0.05 | Balanced, <2 |
|  | PC5 | Contin. | -0.085 | 1.10 | -0.008 | 1.00 | Balanced, <0.05 | Balanced, <2 |
|  | PRS | Contin. | 0.070 | 1.00 | 0.008 | 1.00 | Balanced, <0.05 | Balanced, <2 |

Covariate balance. Diff. = standardised mean difference; VR = variance ratio; unadj. = before weighting; adj. = after weighting. Balance thresholds: Diff < 0.05 for means, VR < 2 for variance

**Supplementary Table S4.**
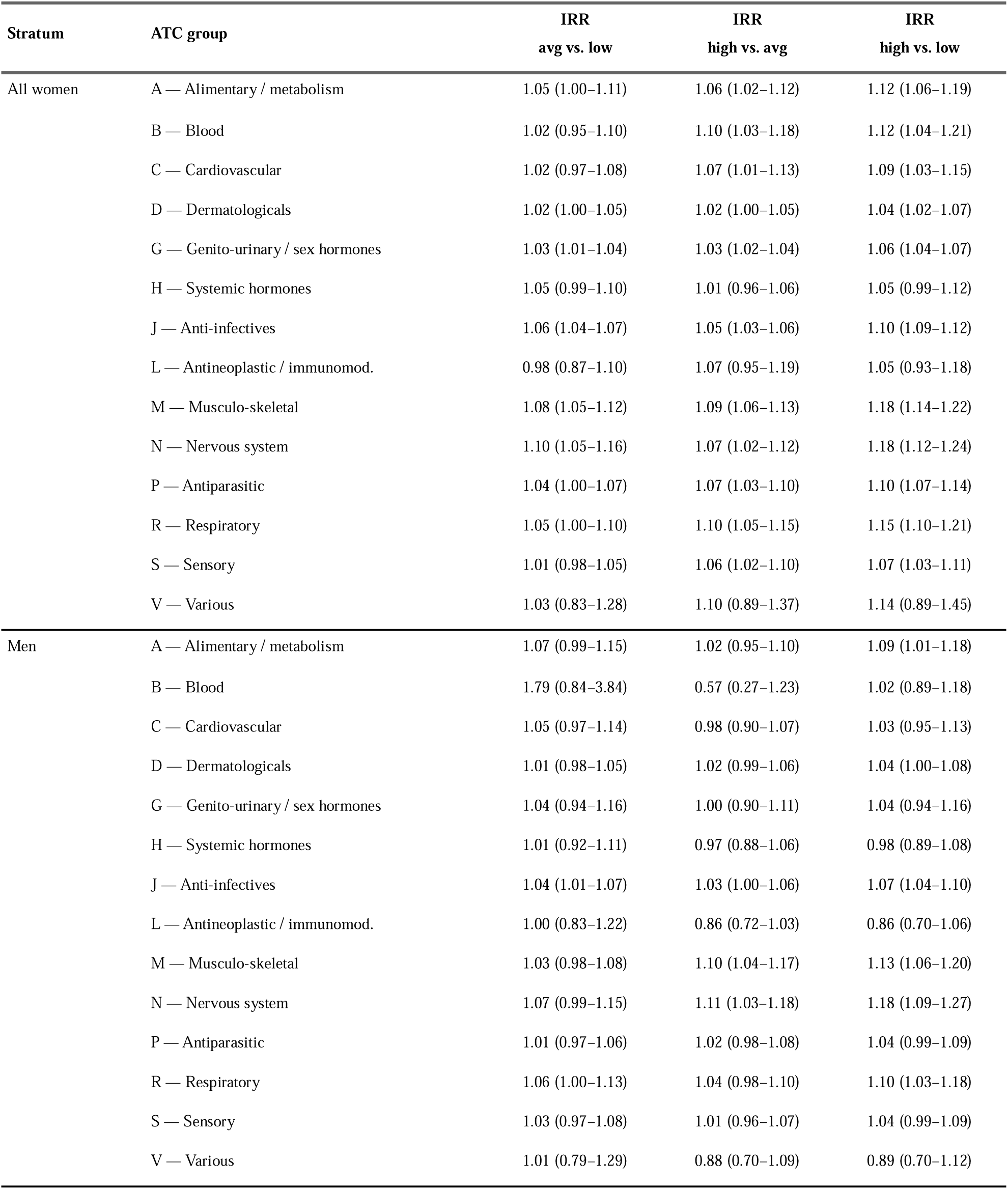
Prescription incidence by ATC anatomical main group, stratified by polygenic risk score (PRS) risk group (low, deciles 1–3; average, deciles 4–7; high, deciles 8–10). Incidence rate ratios (IRR) from quasi-Poisson regression with offset for log observation time, IPTW-weighted. Presented for all women and men.

